# Nowhere to Go: A parallel convergent mixed methods study examining the health of people who experience emergency shelter service restrictions

**DOI:** 10.1101/2024.05.07.24306964

**Authors:** Suraj Bansal, Stephanie Di Pelino, Jammy Pierre, Kathryn Chan, Amanda Lee, Rachel Liu, Olivia Mancini, Avital Pitkas, Fiona Kouyoumdjian, Larkin Lamarche, Robin Lennox, Marcie McIlveen, Timothy O’Shea, Claire Bodkin

## Abstract

**Background:** Emergency shelters offer temporary sleeping accommodation to people deprived of housing and connect them to services. Service restriction is the practice of limiting or denying someone access to emergency shelters. This parallel convergent mixed methods study describes the characteristics, healthcare utilization, and morbidity of people experiencing service restrictions in Hamilton, Ontario, and explores the relationship between health and service restriction.

**Methods:** We recruited 20 people who had experienced service restriction and accessed healthcare from the Shelter Health Network clinic. We conducted semi-structured interviews and performed reflexive thematic analysis. We reviewed participants’ medical records from January 1, 2018 to December 31, 2021 to calculate simple descriptive statistics. Mixing our qualitative and quantitative results, we generated narrative metainferences. We employed community-based research principles, including a research team with lived and living experiences of being service restricted, implementing service restrictions, or providing care to people experiencing service restrictions.

**Results:** We generated six themes: 1) Losing your home shouldn’t mean losing your humanity, 2) Where am I supposed to go?, 3) The snakes and ladders of service restrictions, 4) Abandoned to survive, 5) Constantly criminalized, 6) Harnessing the wisdom of community. Participants averaged 17.4 primary care visits, 11 emergency department visits, and 4 hospital admissions over 4 years. The most common reasons for visit were infections, traumatic injuries, and substance use-related concerns. Narrative metainferences highlighted how people experience dehumanization when accessing shelters or healthcare; how service restrictions and encampment living contribute to infections; the lack of practical supports for people using substances in shelters; the ubiquitous criminalization of people experiencing homelessness; and the care people practice for one another to reduce substance-related harms.

**Conclusions:** Participants’ high healthcare need and utilization was shaped by criminalization, stigma, societal abandonment, and abstinence-based substance use policies. Participants practiced care for themselves and others to navigate these barriers. Shelters should have a transparent service restriction process and employ harm reduction practices. Healthcare should provide affirming and accessible treatment for common conditions. Social and health services must contend with broader social forces while building on the strengths of people with lived experience to improve the health of people who are service restricted.

## Background

Stable housing is an essential determinant of individual and community health. People experiencing homelessness have a mortality rate up to 8 times higher than the general population, with higher rates of cardiovascular disease, mental health disorders, and substance use disorders.^1,2^ This relationship is multi-directional: individuals with poor health are more likely to experience homelessness due to common distal determinants like colonization, ableism, and socioeconomic status; and homelessness itself can cause and exacerbate poor health. Homelessness and mortality are not evenly distributed across the population, but overdetermined by social processes. For example, colonization and anti-Indigenous racism contribute to the overrepresentation of Indigenous peoples among Hamilton’s homeless population at 23%.^3,4^

Emergency shelters provide temporary sleeping accommodation at night and connect users with permanent housing supports. In the city of Hamilton, Ontario, Canada, the 2021 Point-in-Time Count (PiTC) reported 545 people deprived of housing, only 67% of whom used an emergency shelter or city-funded hotel room. These findings indicate that many homeless people in Hamilton experience unsheltered homelessness.^3^ The COVID-19 pandemic caused a national increase in unsheltered homelessness, and put additional strain on Hamilton’s shelter system. A lack of affordable housing, compounded by isolation protocols, disease transmission in shelters, and the release of many of Hamilton’s incarcerated population earlier in the pandemic has contributed to a rise in local homeless encampments.^5,6^

A significant barrier to accessing emergency shelters is service restrictions, the practice of temporarily limiting or denying access to shelters. Available data suggest this is a common practice in Canada; almost 1 in 5 women and gender diverse people surveyed in the 2021 Pan-Canadian Women’s Housing and Homelessness Survey reported experiencing service restriction.^7^ Another survey of 700 Canadian workers in the homeless-serving sector reported that 44.6% had temporarily service-restricted someone in the last month while 14.1% had enforced a permanent ban.^8^

Substance use is common among people deprived of housing, and abstinence-requiring shelter policies pose a significant barrier to people who use drugs (PWUD) accessing shelter services. Hamilton is grappling with a toxic drug supply crisis, as evidenced by 814 suspected overdoses and 167 deaths in 2022 due to opioid-related drug toxicity, surpassing the provincial average.^9^ Prior studies have demonstrated how abstinence-based policies create a high-risk environment for PWUD by forcing people to either avoid shelters altogether, or use substances furtively to avoid surveillance and punishment, ultimately increasing their risk of overdose.^10^ Shelters in Hamilton impose service restrictions for possession of illicit drugs or harm reduction supplies. While designing our study, we received and reviewed shelter standards from two shelter operators in Hamilton; both explicitly stated that possession or use of drugs, alcohol, or drug-use equipment would result in service restriction.

We conceived this exploratory, community-based parallel convergent mixed methods study in response to concerns raised by people experiencing service restrictions and frontline healthcare providers in Hamilton about the detrimental health impacts of service restriction. Despite their pervasiveness, service restrictions are a minimally researched phenomenon. We are aware of one peer-reviewed study on service restrictions that analyzed two different Canadian datasets with logistic regression to identify predictors of service bans.^8^ Kerman et al. determined that victimization and criminalization during homelessness increases risk of service restriction. They identified predictors of service bans that varied between cities within a single dataset, suggesting that organizational, community, and sociopolitical factors are relevant beyond solely individual factors.

To complement the Kerman et al. study, we examine the specific Hamilton context to understand what organizational and community-level factors are related to service restriction. We aimed to understand how service restrictions shape the experiences, healthcare utilization, and morbidity of shelter users experiencing service restrictions in Hamilton, Ontario, Canada.

## Methods

### Study Design, Setting, and Recruitment

We conducted a mixed methods study utilizing a convergent parallel design with people experiencing service restrictions in Hamilton, a midsize city in Ontario, Canada.^11^ We identified 20 participants who met the eligibility criteria of having accessed a homeless shelter and experienced service restrictions and having obtained healthcare via a Shelter Health Network clinic in Hamilton. The Shelter Health Network comprises healthcare professionals partnered with social service organizations to improve health outcomes for individuals who are precariously housed. Participants were recruited through Shelter Health Network clinicians and outreach workers from the research team who were available nearby during street outreach with Keeping Six—Hamilton Harm Reduction Action League, a grassroots drug user advocacy group. Frontline providers would identify potential participants using primarily prior knowledge of whether the individual had experienced service restrictions and received care from the Shelter Health Network. Upon obtaining the individual’s consent, prospective participants were referred to an on-site research team member to enroll if eligible.

We collected qualitative data via semi-structured interviews and quantitative data from participants’ electronic medical records (EMRs). We performed separate qualitative and quantitative analyses by analyzing interview transcripts with reflexive thematic analysis and EMR chart data with simple descriptive statistics.^12^ We then mixed the data by examining the themes and descriptive statistics side-by-side to generate narrative metainferences. We selected a sample size of 20 participants to capture a diversity of experiences with service restrictions and accessing healthcare, while remaining small enough to analyze interviews in depth.

### Data Collection

We developed an interview guide (see Additional File 1) to guide our semi-structured interviews. The interview guide was developed and refined internally and informed by our peer investigator who ensured all questions were respectful and accessible. Interviewers asked participants about their physical and mental health, and lived or living experiences with homelessness and service restrictions. Interviews were conducted between June to August 2022, were 45-60 minutes in length, and were recorded and transcribed verbatim. Participants received refreshments and food, and were compensated with a $30 cash honorarium in line with the British Columbia Centre for Disease Control guidelines for peer payments.^13^ We developed and piloted a data extraction template to uniformly collect data across all charts (available upon request). We extracted quantitative data from the Shelter Health Network EMR, Hamilton Health Sciences EMR Meditech, and St. Joseph’s Healthcare Hamilton EMR Dovetale for all study participants. The data contained both sociodemographic data and health outcomes from the study period of January 1, 2018 to December 31, 2021.

### Quantitative Data Analysis

Quantitative data included demographic data, and information on participants’ interactions with the healthcare system, including primary care visits and diagnoses, emergency room visits and diagnoses, and hospital admissions and diagnoses.^14^ To reduce the overrepresentation of individuals using primary care services more frequently, we limited chart review and data extraction to the first 25 visits for each participant. We prioritized the extracted data for downstream analysis using both existing literature on morbidity in people experiencing homelessness, and insights from co-investigators based on frontline experiences with patients. We also categorized data according to ICD-10-CA codes, where applicable.^1,2^ Using descriptive statistics, we reported the number of visits, reason for visits, housing status, and disposition destination.

### Qualitative Data Analysis

Interviews were transcribed, coded, and analyzed using reflexive thematic analysis.^15^ Several research team members coded interview transcripts independently using the Dedoose software and met regularly to review codes, develop an inductive coding framework, explore and resolve discrepancies, and generate themes.^16^ We continued to refine themes while extracting the supportive quotes from participant interviews. We championed a social justice and harm reduction orientation stemming from the community-based organizing and political work done by several members of our research team. Moreover, we brought our lived and living experiences of housing deprivation and being service restricted, implementing service restrictions, and providing care to people experiencing service restrictions, to our reflexive thematic analysis, reinforcing the validity of our findings.^17^

### Mixed Methods Analysis

Themes and descriptive statistics were examined side-by-side to understand how they complement, elaborate, and contradict each other. We then generated metainferences in the form of narrative statements, framed by our assumptions and worldviews as stated above.^18^

### Community Based Research Principles

Since our study concerned people in our community, living and experiencing multiple forms of oppression and marginalization, we employed several community-based principles to ensure our research contributed to changing community conditions. Notably, our research team included a peer investigator with lived experience of homelessness and service restriction, someone with frontline experience enforcing service restrictions in shelters, and several co-investigators with frontline community-based clinical experience providing care to people facing service restrictions. The peer investigator helped ensure our research questions, methods, and knowledge translation activities promoted the dignity, autonomy, and wellbeing of people experiencing homelessness and service restrictions.^13,19^ We hosted research team meetings in person and provided virtual participation options; provided food at all meetings; and compensated transportation for peer investigators. To uphold our accountability to the community, we hosted a barbeque at a large homeless encampment to disseminate our results, and held a stakeholder workshop to promote uptake of our findings among people with decision-making authority.

### Critical Analytic Frameworks

Throughout this study, our research team drew on two critical frameworks. Through Eve Tuck’s concept of Suspending Damage, we went beyond merely documenting “peoples’ pain and brokenness to hold those in power responsible for their oppression,” by considering how people facing service restriction are navigating their own ways to improve individual and community health.^20^ Embracing this approach helped draw out the numerous strengths participants expressed about themselves and their communities during interviews, turning our attention towards the possibilities for community-led change.

The Latin American concept of Social Determination of Health helped us understand how structures and systems such as racism, criminalization, and capitalism all intersect to undermine the health of people who are service restricted; while collective care organized by PWUD to advance harm reduction resists and reshapes some of these processes.^22^ While social determinants of health overemphasize discrete factors and linear unidirectional causal pathways, social determination of health recognizes that many intersecting sociopolitical and ecological forces create the processes and conditions that shape and are in turn shaped by collective and individual health.^21^ Furthermore, social determination of health acknowledges that not all of these processes or conditions are empirically measurable and requires us to embrace other modes of inquiry to understand them. Mixed methods analysis enabled us to measure the high prevalence of illness in people experiencing service restrictions and unpack how sociopolitical forces within and outside service restrictions shape individual and community health.

## Results

We developed six themes in our analysis. The themes are interrelated and presented in no particular order:

1. Losing your home shouldn’t mean losing your humanity
2. Where am I supposed to go?
3. The snakes and ladders of service restriction
4. Abandoned to survive
5. Constantly criminalized
6. Harnessing the wisdom of community

### 1. Losing your home shouldn’t mean losing your humanity

Participants’ humanity is threatened while experiencing homelessness. Shelter users often described feeling labeled, dehumanized, and treated without compassion when discussing their interactions with shelter staff. Some participants identified an apparent lack of staffing, training, and sensitivity towards those accessing shelters.

> “Then we’re no longer looked at as people. When you’re not dealing with human life, we’re not looked at as people and you’re not dealing with life itself, you’re now just a number, or just an addiction problem, or a mental health problem, or a disability.” (Interviewee 9)

Similar feelings of dehumanization are felt among participants when accessing healthcare, with some PWUD facing stigma from healthcare professionals. As a direct result of being treated differently based on their substance use, people avoid seeking healthcare.

> “Instantly the nurses absolutely are judgmental…we have an addiction, yes. Sorry. But you know, like we’re not animals, we’re not monsters. You know, like we say, you don’t know what we’ve been through. So, yeah, they’ve got to stop being so judgmental” (Interviewee 7)
>
> “If you’re homeless they just want to rush you through as quick as they can and get you out the door. They don’t want you in there. You’re in there and clogging up their beds and clogging up the system, so they just try to push you out the door.” (Interviewee 6)

Participants expressed that their physical and psychological safety is constantly challenged and threatened. There is a perpetual cycle of trauma from the constant loss and theft of belongings while homeless or following a service restriction. These physical conditions can cause or exacerbate existing mental health, physical health, and substance use related conditions for some shelter users.

> “Affected my mental health more than anything else because you lose everything you have when you’re service restricted. They steal it all. Whatever is there they throw it away somehow and then say they lost it.” (Interviewee 10).
>
> “I’m left with a very strange feeling at night when you’re not housed, eh. You don’t know who’s coming down the street. You don’t know what’s going to happen” (Interviewee 8)
>
> “I get anxiety all the time because, like, it’s stressful when you don’t have a place to go so you’re—like for the day, you’re stressing out about where you’re going to sleep at night.” (Interviewee 2)

Despite these physical and psychological conditions experienced while living on the streets, numerous participants feel safer there than in shelters. Participants describe a lack of privacy, and a violent jail-like environment that make shelters a stressful and uncomfortable place to access. Shelter residents feel like they are constantly negotiating how to meet their needs and spend their time under surveillance of staff. There was a common undertone that human connection is the foundation of feeling a sense of home, which is lacking in the current shelter system.

> “I didn’t find shelters any less stressful than just being on the street, because it’s the same type of people.” (Interviewee 19)
>
> “And people steal and people don’t steal, and people go without things because people take things away from them, you know. It’s just kind of hard to start your life over until you’ve actually moved into a new home, you know.” (Interviewee 8)
>
> “Without [human contact], taking away the essential need for people to communicate and feel empathy for each other, or help share experiences, makes the experience lonely, sad, angry and eventually explosive.” (Interviewee 10)

Ultimately, study participants wanted a home, not just a shelter bed.

> “Like, maybe I’m tired of moving. I don’t want to move all the time. I really don’t want to start over again. You know, well, I mean I will have to start over, but I don’t want to have to do it all the time, you know what I mean. I want to move somewhere and stay there.” (Interviewee 8)

Homelessness removed people from the bureaucratic systems of society and created many intersecting legal barriers to re-entering the workforce. This included not having a bank account, lacking a fixed address, or potentially having a criminal record. PWUD also expressed concern about stigma towards them if they wanted to enter the workforce, and the distress that current substance use could impact their performance in the workplace.

> “No bank account, no e-transfers. You’re less of a person. Unable to rent a vehicle, lease a vehicle—less a person, you know. Criminal record—you can’t own a City registered business.” (Interviewee 12)

### 2. Where am I supposed to go?

Being service restricted is more than just losing a bed. Following service restriction, participants are cut off from social and health services. For some, being service restricted increased barriers to accessing healthcare and increased their use of the emergency department. This was enacted by rules that limited access (e.g. being service restricted denied people from accessing on-site medical care). Furthermore, even if they were permitted to access services like medical care from shelters they were previously banned from, the distance from their new shelter made it infeasible. The internalized and external stigma following service restriction also became a barrier to accessing essential services.

> “So any service they could forward, it sounds like they’re restricting you, and they’re not giving you access to any of it. So I mean to me, it sounds like they’re shutting them down all across the board.” (Interviewee 4)
>
> “If you get service restricted from somewhere where, you know, it’s comfortable for you, and then the only place you can go is somewhere where you’re not comfortable going, you’re basically guaranteed to be outside.” (Interviewee 12)
>
> “I was too embarrassed to go back and collect my things and find out if I can either get my methadone there, or whether I have to go back to the clinic and drop from a high dose that helps out a bit, to a low dose that I almost won’t feel at all.” (Interviewee 12)

With nowhere to go, people generally either joined encampments or floated around outside. Participants described how service restrictions directly contribute to the growth of encampments.

> “I just gave up and I just pitched a tent and became a part of that community.” (Interviewee 11)
>
> “[service restriction] causes it to grow, and living out is better than nowhere.” (Interviewee 12)

The lack of access to basic necessities in encampments worsened health outcomes of encampment residents.

> “Health was horrible. Every single person I came in contact with had some type of infection, sickness. And it was just—yeah, it was just running rife throughout the camps.” (Interviewee 11)

Winter was identified as a particularly important yet difficult time to be able to access shelter.

> “Then I found myself trying to find a stairwell to stay in, because it was freezing. It was during the winter. So I ended running and sneaking into the Sheraton, downstairs into the basement parking lot, running to a stairwell before the security guards caught me … I woke up to cops giving me a ticket, a $70 ticket.” (Interviewee 19)

### 3. The snakes and ladders of service restriction

There was a common experience among participants that the process and rules of service restriction were arbitrary and unjust. Participants felt rules were applied at random depending on the individual shelter staff enforcing them. This contributed to the growing mistrust between shelter staff and users, and a common feeling among participants that staff misused their authority. Participants felt that rules were unrealistic, and it was impossible to abide by them especially if you are a person who uses drugs.

> “There’s like women out there that do sex work, and they would lose their bed because they weren’t home, because they had to make their money at night, which makes it really hard for that person.” (Interviewee 13)

There is a need for transparency regarding service restrictions and appropriate processes for appeal and mediation. It was suggested that conflict resolution, mediation, and meaningful involvement of those with lived or living experiences could be ways to improve the current service restrictions processes.

> “Offering an appeal process. Being clear on the reasons why, and showing the rules that were broken … be clear, give warnings, have a protocol for it that they have to follow. A couple of warnings. And the warnings themselves can be appealed. Because I’ve been tricked by that too and kicked out. And they just say that they warned you about something that they didn’t warn you about.” (Interviewee 10)
>
> “To have somebody who’s not working with them and working for them, you’ve got to have somebody in the middle, you know, like a mediator, who’s going to be fair and not working with the police and not working with the system and not working with any of them; just a regular person who could, you know, empathize, sympathize with people, and you know. That would help a lot. I think that would make a big difference, man. That would be it. And not have them make that decision, that final call, you know.” (Interviewee 11)

Substance use was a common reason for service restrictions, and attempting to avoid service restriction created a heightened risk environment for PWUD.^15^ Abstinence-based rules for substance use in shelters forced PWUD to choose between either avoiding shelters altogether and staying outside or using shelters and trying to avoid service restriction by hiding their substance use. For some participants, service restriction forced them back to the streets and precipitated relapse to substance use or increased substance use.

> “Like, I guess being more understanding or like lenient when it comes to people getting service restricted for reasons related to their addiction and stuff.” (Interviewee 2)
>
> “I’m gonna go out there. It’s cold. I’m gonna probably end up in an abandoned building, and there’s gonna be dope everywhere and I’m probably gonna relapse. I hope you feel good about yourself. … this one [shelter staff], she’d done it to three different people.
>
> And my one friend relapsed using alcohol, this other one relapsed using crystal meth; all for the same reason because this woman was on a high horse and kicked us out for nothing, just stupid, childish things.” (Interviewee 7)

### 4. Abandoned to survive

Most participants described their ongoing struggle to fulfill essential material needs required for maintaining good health. This challenge persists both within shelters and when facing service restrictions, without family or community relationships to anchor them. There is sustained sleep deprivation both in shelters and outside, insufficient access to high-quality food, low income support payments, high market rents, and minimal subsidized or supportive housing.

> “How the fuck am I going to get housed when I can’t even afford the rent?” (Interviewee 9)

There are common narratives of feeling unsupported when coping with acute and chronic pain, as well as mental and physical illnesses. Pain emerged as a significant health concern for numerous participants, often leading to substance use. For some, substance use felt like the only option, stemming from inadequate mental and physical health services, and insufficient outreach that meets people where they are at.

> “And now I’m a Fentanyl user because of the pain that I go through with my leg and my hip, everything, because I got to offset it. And I was told I can’t get a; I’m not going to get a [doctor’s] appointment because I’m not housed.” (Interviewee 9)
>
> “Self-medicating is really the only way to maintain life as a homeless individual, period. And really that’s all it is. And substance abuse, I don’t even think should be the term used, I think it should just be self-medicating. That’s all it is ever is. Nobody abuses substances. They medicate themselves to the point that they feel comfortable again with whatever they have access to. So, if you got pills from a doctor, drugs on the street or grass out of a field, it doesn’t really matter what works. Whatever works for someone they’re going to use.” (Interviewee 10)

Since current shelter policies prohibit people from possessing or using drugs, and noting that participants were recruited partially via Keeping Six, many participants spoke to their experiences of either avoiding shelters or being service restricted due to substance use. Everyone acknowledged widespread substance use in shelters presently.

> “You can’t use inside—you can’t—that’s their rules. You can’t use on property.” (interviewee 12)

Many participants identified loss and breakdown of family relationships as the root of instability in their lives. Participants explained how this negatively impacted their mental health and contributed to escalating substance use and becoming homeless.

> “When I lost my son, that’s when I got hard into drugs” (Interviewee 7)
>
> “The way I ended up out on the street was a real bad ordeal. I was told I wasn’t going to get my kids back. I had full custody of my kids. And I was told I wasn’t going to get my kids back after I had jumped through the fucking hoops like the system wanted me, to and then some.” (Interviewee 9)

### 5. Constantly Criminalized

Although the interview guide did not include any questions about criminalization or incarceration, almost every participant identified that policing, incarceration, and criminalization affected their lives and health. These interactions with the justice system often involved inappropriate or threatening use of police authority during instances of service restriction.

> “And so then I was arrested. And they made sure that I wasn’t getting out. They tried to make sure that I got whatever. That shelter made sure, right.” (Interviewee 9)

Some participants mentioned that being service restricted consequently forced them into public areas where they felt constantly surveilled by police, placing them at a higher risk of being stopped or arrested. Police, parole, and bail conditions may also restrict people from certain geographic areas, making it even harder to access essential services when facing service restrictions.

> “Like, my probation doesn’t allow me to go 50 meters near that [name] building across from the [Shelter]. So I don’t want to go to the [Shelter], I’m going to jail, you know what I mean. I’m only permitted there between 10:00 and 1:00 o’clock, you know what I mean. So, I’m basically crossing an imaginary line and every time I do and I’m caught, I’m being thrown in jail for it, you know what I mean.” (Interviewee 8)

There was also frequent police presence when accessing healthcare. Several participants reported avoiding seeking care due to police presence in these settings. Others described this in a matter-of-fact way, describing it as a feature of accessing healthcare in hospitals or via emergency medical services (EMS).

> “The whole time I was in the hospital, I had like an orange jumpsuit and I had to walk around cuffed and shackled with two guards, one on each side of me, the whole time, anywhere I walked.” (Interviewee 2)

### 6. Harnessing the wisdom of community

People have many ideas to improve shelter systems, informed by their first-hand experiences of its shortcomings. Participants called for a transparent, consistent process for service restrictions that includes timely appeals and neutral third-party involvement. They highlighted the need for practical substance use policies, more privacy in shelters, and shelters tailored to different demographics, such as older adults or couples without children. Additionally, participants underscored the importance of providing immediate alternatives when service restricting someone, instead of having them locate it themselves.

> “They should have like a intermediate person, like someone, like a mediator, right; like someone in the middle, to really lead, like, not bias and stuff and really look at the whole situation and make a decision then; not just have a person say, “You did this, so you’re done. That’s it. No discussion. You’re out of here.” You know what I mean? Because it’s heartbreaking, man, you know, guys getting restricted for stupid things, and that’s it, there’s no option, they’re just, they’re done. And it’s pretty annoying” (Interviewee 7)
>
> “There should always be somewhere where you can go, you know. It shouldn’t be totally—in a city of this size, there shouldn’t be a person that’s totally restricted from everywhere. It’s minus 40 in the winter.” (Interviewee 12)

Many participants were involved in harm reduction both informally (e.g. checking on friends using drugs or acquiring and distributing sterile drug use supplies) and formally (e.g. volunteering with Keeping Six). The social determination of health framework invited us to consider how this community knowledge and action serves to resist and reduce harm from both Canada’s and shelters’ prohibitionist drug policies.

> “I’m involved with harm reduction because I believe it saves lives, the people… Yeah, like I had a bunch; a package, a ten-pack, a two-pack, whatever, and anybody that required them, I would just hand them out to them, because that’s what I do.” (Interviewee 1)

Participants called for alternatives that enable PWUD to safely and reliably access shelters, including revised abstinence-based rules that acknowledge the reality of substance use and permit shelter users to possess and use drugs. People described the safety improvements they saw in shelters with designated drug use spaces and suggested that such spaces be widely implemented. A few participants proposed the establishment of both sober and wet shelters to acknowledge that staying abstinent to substance use is challenging given its existing pervasiveness in shelters.

> “This one has one, has a safe injection site in it and it’s been a lot better over there for a lot of the females than it was before.” (Interviewee 17)
>
> “I think every place should have a safe site for people that use.” (Interviewee 16)

Study participants reiterated long-standing calls for healthcare to be non-judgemental, accessible, and meet people where they are at. This contrasted their lived experiences in healthcare settings, particularly in hospitals. Participants noted the importance of street outreach conducted by clinics and physicians. While physical healthcare was more accessible, most participants highlighted the lack of accessible mental health resources, including psychiatric care, counseling, residential treatment, social workers, and other non-pharmacologic therapies. Some positive healthcare experiences were mentioned, but these were considered rare exceptions. Access to medication was generally reliable. Many participants also expressed the importance of the Shelter Health Network in providing accessible and reliable healthcare.

> “Just do outreach. More outreach and more and more showing up where people are at instead of having them to come to you, right.” (Interviewee 6)
>
> “They do a lot. I’ve watched what they do. They’ve done everything, from giving tents, to shoes, clothing, you know, medical aid, you know… I told the doctor, I said, “I’m getting healthy because a lot of good people are helping me,” (Interviewee 8)

Despite the hardships, challenges, and mistreatment, there was a resounding sense of community solidarity among those surviving homelessness and service restrictions together. Participants described themselves as compassionate and capable of coping as a community with hardships, despite a system working against them.

> “I still help people and give the shirt off my own back, and I’m homeless, and have nothing. And somebody that has less than what I have, I still give. And that’s what most people ever give.” (Interviewee 8)

### Quantitative Results — Descriptive Statistics

Participants’ ages ranged from 26 to 69 years, with an average age of 42.9 years. 30% of participants self-identified as Indigenous, 5% as Black, and 55% as White. 75% of participants self-identified as men. The sample was recruited through convenience sampling, and we suspected that individuals who have experienced service restrictions have distinct characteristics compared to the general homeless population. Compared to the 2021 PiTC, the study sample was similar in age and representation of Indigenous people, but contained a lower percentage of Black people and women.^3^

**Table 1:** Characteristics of study participants who had experienced service restriction in Hamilton, Ontario, N=20.

|  |  |  |
| --- | --- | --- |
| **Age- years** | Mean | 42.9 |
|  | Minimum | 26 |
|  | Maximum | 69 |
| **Gender\*- n (%)** | Man | 15 (75) |
|  | Woman | 4 (20) |
|  | Transgender Woman | 1 (5) |
| **Sexual Orientation\*- n (%)** | Straight | 17 (85) |
|  | Asexual | 1 (5) |
|  | Lesbian | 1 (5) |
|  | Bisexual | 1 (5) |
| **Racial Identity\*,- n (%)** | Métis | 2 (10) |
|  | First Nations | 4 (20) |
|  | White | 11 (55) |
|  | Black | 1 (5) |
|  | Mixed Race | 3 (3) |
| **Income\*- n(%)** | Ontario Disability Support Program (ODSP) | 12 (60) |
|  | Pension Income | 1 (5) |
|  | Ontario Works | 4 (20) |
|  | Informal economies** | 13 (65) |
|  | No Income | 1 (5) |

\* Self-identified
+ Does not add to 100% as some participants identified with more than one category
\*\* Panhandling, odd jobs, casual jobs, casual peer outreach, street hustling, sex work

The study participants frequently used primary care services, averaging 17.4 visits per person. Housing status was not routinely recorded in the medical charts, with 67% of visits lacking documentation of the person’s housing situation at the time of visit. Wounds, substance use, and infections were the most common reasons for seeking primary care. Many visits were also devoted to accessing social services (e.g. completing ODSP applications). Notably, while primary care typically strives to provide preventative care and chronic disease management, only 5.1% of visits were dedicated to this.

**Table 2:** Primary Care Visits from 2018 - 2021 for people who experienced service restriction in Hamilton, Ontario.

| **Primary Care Visits (N = 330 visits, representing 19 study participants)** |  |  |
| --- | --- | --- |
| **Description** |  | **Number of Visits** |
| Average number of visits per person |  | 17.4 |
| Minimum visits per person |  | 3 |
| Maximum visits per person |  | Over 45 |
| **Description** |  | **Number of Participants** |
| 0 - 4 visits in study period |  | 2 |
| 5 - 12 visits in study period |  | 7 |
| 13 - 20 visits in study period |  | 4 |
| 21+ visits in study period |  | 6 |
| **Housing status at time of visit (N=279 visits\*)** |  | **N (%\*)** |
| Housed |  | 13 (4.7) |
| Unsheltered homelessness |  | 30 (10.8) |
| Sheltered homelessness |  | 46 (16.5) |
| Not documented |  | 189 (67.7) |
| Restricted from all shelters |  | 1 (0.4) |
| **Reason for Visit (N=279 visits\*)** |  |  |
| **Category** | **Sub-Category** | **N (%\*)** |
| Infections |  | 53 (13.5) |
|  | Abscesses | 9 (2.3) |
|  | Endocarditis | 3 (0.8) |
|  | Osteomyelitis | 8 (2.0) |
|  | Other infections | 33 (8.4) |
| Substance Use Disorder |  | 52 (13.2) |
| Wounds |  | 44 (11.2) |
| Medications |  | 40 (10.2) |
| Mental Health |  | 34 (8.7) |
| Assistance with accessing social services |  | 32 (8.1) |
| Pain |  | 28 (7.1) |
| Other |  | 23 (5.9) |
| Inflammation |  | 18 (4.6) |
| Supportive Visits |  | 15 (3.8) |
| Traumatic Injuries |  | 14 (3.6) |
| Preventative Health |  | 11 (2.8) |
| Headache |  | 6 (1.5) |
| Dental Issues |  | 6 (1.5) |
| Type 2 Diabetes |  | 5 (1.3) |
| Gastrointestinal Issues |  | 4 (1.0) |
| Hepatitis C |  | 4 (1.0) |
| Shortness of Breath |  | 2 (0.5) |
| Respiratory Issues |  | 2 (0.5) |

\*Representing a maximum of 25 visits reviewed per study participant
\*Does not add to 100% due to rounding

Study participants also frequently visited the emergency department, averaging 11 visits per person over study period. In 10% of visits, participants presented to the emergency department while in custody of police or correctional officers. In 14% of visits, patients initiated discharge either before or after consultation with an emergency room physician. Infections, traumatic injuries, and substance use marked the most common reasons for visits, collectively accounting for 65% of visits.

**Table 3:** Emergency Department Visits from 2018 - 2021 for people who experienced service restriction in Hamilton, Ontario.

| Emergency Department Visits (N = 217 visits, representing 19 study participants) |  |
| --- | --- |
| Description | Number of Visits |
| Average number of visits per person | 10.95 |
| Minimum visits per person | 2 |
| Maximum visits per person | Over 27 |

| Description |  | Number of Participants |
| --- | --- | --- |
| 0 - 4 visits in study period |  | 3 |
| 5 - 12 visits in study period |  | 10 |
| 13 - 20 visits in study period |  | 2 |
| 21+ visits in study period |  | 4 |
| Presented From |  | N (%+) |
| Walk-In |  | 121 9 (55.8) |
| EMS |  | 74 (34.1) |
| Brought in by police or correctional officers |  | 22 (10.0) |
| Disposition |  | N (%+) |
| Discharged home |  | 149 (68.7) |
| Admitted to hospital |  | 18 (18.0) |
| Discharged to custody of police or correctional officers |  | 14 (6.5) |
| Discharged to shelter |  | 6 (2.8) |
| Patient-initiated discharge after being seen by ERP* |  | 26 (12.0) |
| Patient-initiated discharge before being seen ERP* |  | 4 (1.8) |
| Reason for Visit |  |  |
| Category | Sub-Category | N (%+) |
| Substance Related |  | 70 (32.3) |
|  | Substance Overdose | 33 (15.2) |
|  | Substance Abuse | 24 (11.1) |
|  | Seeking Detox | 5 (2.3) |
|  | Withdrawal | 8 (3.7) |
| Infections |  | 41 (18.9) |
|  | Abscesses | 16 (2.8) |
|  | Skin and Soft Tissue Infections | 5 (7.4) |
|  | Endocarditis | 2 (0.9) |
|  | Osteomyelitis | 2 (0.9) |
|  | Pneumonia | 5 (2.3) |
|  | MRSA (location/type not specified) | 5 (2.3) |
|  | Other infections | 5 (2.3) |
| Traumatic Injuries |  | 30 (13.8) |
| Pain |  | 21 (9.7) |
| Mood Disturbances |  | 14 (6.5) |
|  | Depression | 3 (1.1) |
|  | Suicidal Ideation | 5 (2.3) |
|  | Behavioral Issues | 6 (2.8) |
| Dental Issues |  | 10 (4.6) |
| Inflammation |  | 10 (4.6) |
| Altered LOC |  | 9 (4.2) |
| Wounds |  | 8 (3.7) |
| Hyperglycemia |  | 5 (2.3) |
| Headache |  | 4 (1.8) |
| Constipation |  | 1 (0.5) |
| Nausea/Vomiting |  | 1 (0.5) |
| Seizure |  | 1 (0.5) |
| Shortness of Breath |  | 1 (0.5) |

\*ERP = Emergency Room Physician
\*Does not add to 100% due to rounding

16% of emergency department visits resulted in hospital admission, with 14% leading to patient-initiated discharge. Substance-related issues accounted for only 5.7% of admissions, which represents less than 3% of all substance-related emergency department visits. The most common reasons for admission were traumatic injuries and infections.

**Table 4:**
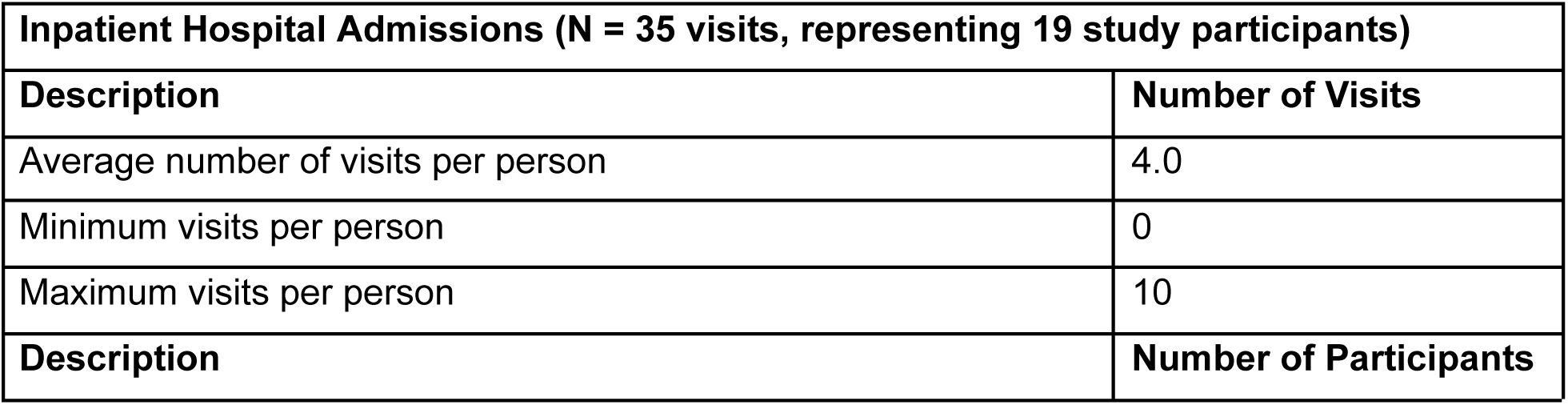
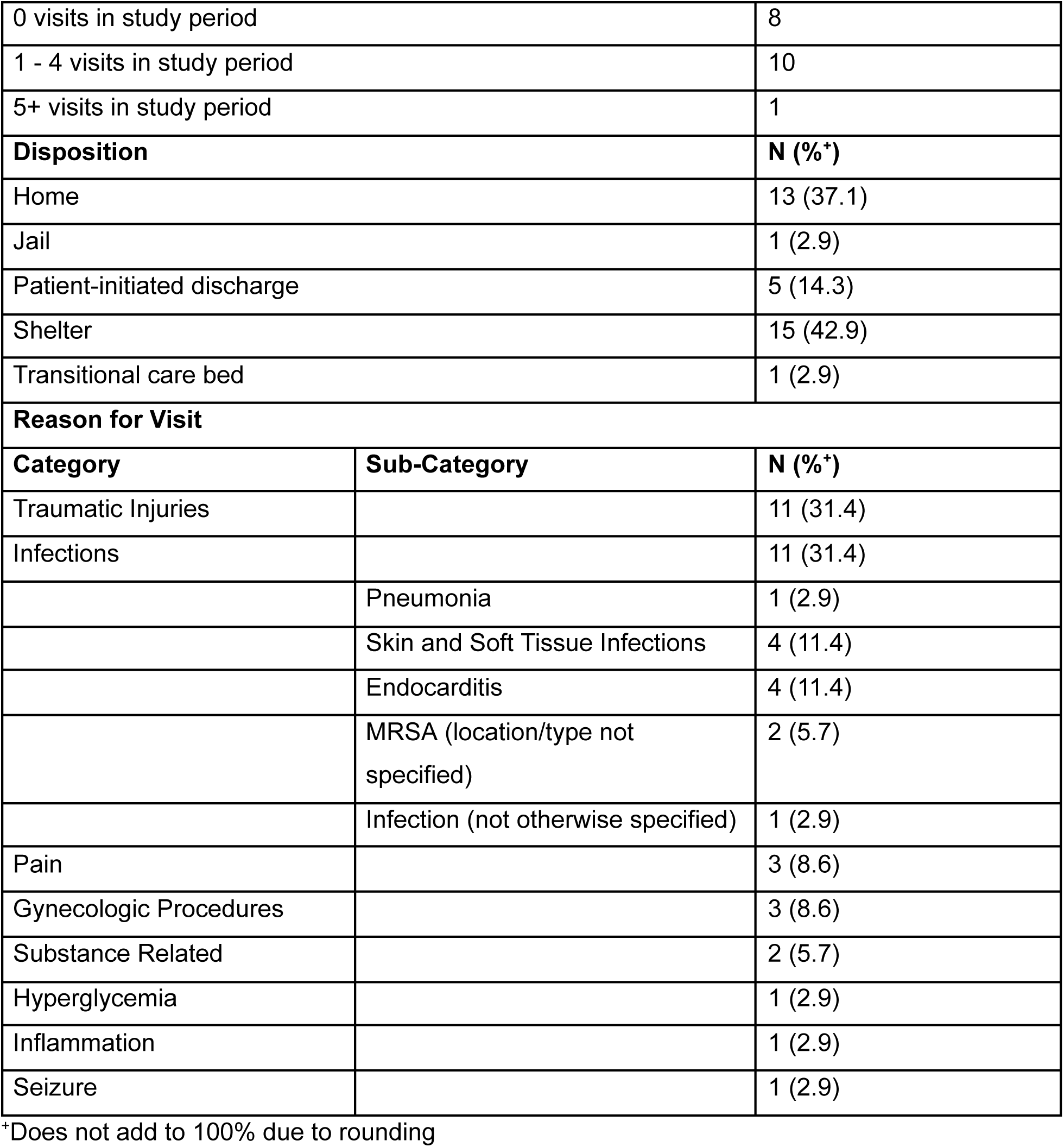
Inpatient Hospital Admissions 2018-2021 for people who experienced service restriction in Hamilton, Ontario.

| Inpatient Hospital Admissions (N = 35 visits, representing 19 study participants) |  |
| --- | --- |
| Description | Number of Visits |
| Average number of visits per person | 4.0 |
| Minimum visits per person | 0 |
| Maximum visits per person | 10 |
| Description | Number of Participants |

|  |  |  |
| --- | --- | --- |
| 0 visits in study period |  | 8 |
| 1 - 4 visits in study period |  | 10 |
| 5+ visits in study period |  | 1 |
| Disposition |  | N (%<sup>+</sup>) |
| Home |  | 13 (37.1) |
| Jail |  | 1 (2.9) |
| Patient-initiated discharge |  | 5 (14.3) |
| Shelter |  | 15 (42.9) |
| Transitional care bed |  | 1 (2.9) |
| Reason for Visit |  |  |
| Category | Sub-Category | N (%<sup>+</sup>) |
| Traumatic Injuries |  | 11 (31.4) |
| Infections |  | 11 (31.4) |
|  | Pneumonia | 1 (2.9) |
|  | Skin and Soft Tissue Infections | 4 (11.4) |
|  | Endocarditis | 4 (11.4) |
|  | MRSA (location/type not specified) | 2 (5.7) |
|  | Infection (not otherwise specified) | 1 (2.9) |
| Pain |  | 3 (8.6) |
| Gynecologic Procedures |  | 3 (8.6) |
| Substance Related |  | 2 (5.7) |
| Hyperglycemia |  | 1 (2.9) |
| Inflammation |  | 1 (2.9) |
| Seizure |  | 1 (2.9) |

\*Does not add to 100% due to rounding

### Mixed Methods Results – Metainferences

“Losing your home shouldn’t mean losing your humanity” (theme 1) highlighted the stigma that people deprived of housing and PWUD face when accessing healthcare in hospital settings. Combined with the pressing survival needs detailed in “Abandoned to Survive”, this may contribute to high rates of self-initiated discharge from emergency departments and inpatient hospital admissions. Traumatic injuries were a common reason for emergency department visits and inpatient hospital admissions, and were reflected in the lack of physical and psychological safety reported by participants.

“Where am I supposed to go?” (theme 2) articulated the pathway from service restriction to joining an encampment, and the resulting health challenges like contracting an infection without access to basic hygiene facilities. This was mirrored in the high frequency of visits for infections in primary care, emergency departments, and inpatient admissions. Although several participants were reluctant to seek hospital-based healthcare, echoed in themes of “Where am I supposed to go,” “Abandoned to survive,” and “Losing your home shouldn’t mean losing your humanity,” they still represent a group with high healthcare utilization. This was evidenced by the significant number of visits across all three care settings included in our study.

“Abandoned to survive” (theme 4) highlighted the lack of access to timely mental health and substance use care, while “Snakes and ladders of service restriction” (theme 3) described how abstinence-based policies in shelters compromised PWUDs’ ability to access shelters. Rather, PWUD were forced to hide substance use in spaces like bathrooms, often resulting in service restriction, or avoid shelters altogether. This was reflected in a high frequency of emergency department visits for substance use issues. Although hospitals may not be the most appropriate or desirable setting to receive substance use care, low admission rates after presentation with substance use issues suggests that people seeking care are not offered inpatient admission in instances where it may actually benefit them.

While criminalization was not our study’s initial focus, it constantly resurfaced in both quantitative and qualitative analysis. We noticed a multidirectional relationship between housing deprivation, service restrictions, and healthcare access. “Constantly criminalized” (theme 5) spoke to avoiding hospital-based care settings to avoid possible police interactions or arrest. Criminalization is a tool to enforce service restriction, a deterrent from seeking healthcare, and a common experience in everyday life.

Finally, “Harnessing the wisdom of community” (theme 6) revealed that study participants practice care to others experiencing the harms of the contaminated drug supply. They call for pragmatic alternatives to abstinence-based shelter policies such as safer use spaces in shelters, and separate shelters for people who are sober or using substances. This responds to the high rates of infection reported in our study by promoting safe injection practices to minimize injection-related infections. It may also reduce the frequency of substance-use related emergency incidents by providing people with a supervised setting where drug usage is not rushed and overdoses can be managed onsite without requiring EMS or hospital transfers. Participants also called for being treated with dignity and humanity in healthcare settings, which may address the high rates of patient-initiated discharge.

## Discussion

Our research reinforces Kerman et al.’s prior finding that victimization and criminalization during homelessness may increase risk of service restriction.^8^ While Kerman et al. identified that service bans are predominantly shaped by organizational, societal, and community factors rather than individual circumstances, we uncovered what these organizational and community level factors are.

Participants found service restrictions opaque and inconsistent, calling for service restriction policies to be transparent and accountable with an accessible appeal process. They recommended a pragmatic approach to substance use in shelters to reduce the frequency of service restriction and substance-related harms among people accessing emergency shelters. Participants were neglected by systems and society while trying to survive, yet discovered strength and support in caring for themselves and one another. Embracing Eve Tuck’s concept of Suspending Damage, we went beyond merely documenting “peoples’ pain and brokenness to hold those in power accountable for their oppression” by considering how people facing service restrictions are navigating their own ways to improve individual and community health.^20^ As a direct consequence of experiencing its shortcomings firsthand, they developed several ideas to improve the shelter system. Participants valued and practiced harm reduction strategies to safeguard against harms associated with the contaminated drug supply and abstinence-based shelter policies.

The Latin American concept of Social Determination of Health offers a framework for understanding how the structures of policing and criminalization underpin homelessness and health disparities.^21, 22^ Despite no interview questions directly addressing criminalization, many participants reported that threats of police action and incarceration were used to implement service restrictions, ultimately deterring them from accessing healthcare. Following service restriction, individuals are displaced into public spaces where they experience increased surveillance and police targeting. Ubiquitous threats of police detention and arrest, particularly in hospital settings, deters people from accessing healthcare. This further intersects with oppressive forces like racism, colonization, and ableism that determine who is over-policed, over-incarcerated, and overlooked by our healthcare system.

### Study Limitations

The study sample is not representative of all people experiencing service restrictions in Hamilton. We recruited participants through outreach alongside Keeping Six, a community organization led by and serving PWUW, and were therefore more likely to recruit PWUD. There were high mortality rates for PWUD throughout the study period and given our recruitment methods and the retrospective chart review, this study only includes people who were alive at the end of the study period. We cannot speak to the unique experiences and health conditions of PWUD who were service restricted and died during the study period. Consequently, our quantitative work is more illustrative and exploratory. However some of the findings may be generalizable. Although the exact number of people experiencing service restrictions in Hamilton each year is unknown, we estimate that our sample represents approximately 5% of those facing service restrictions from Hamilton shelters. This estimate is based on data about homelessness in Hamilton and the prevalence of service restrictions reported in recent national surveys.^3,7^ Despite asking what communities our participants identified with and grounding our themes in the social determination of health, our qualitative data’s thickness was insufficient to adequately explore the effects of race, colonization, gender, or sexual orientation in our themes. Furthermore, without access to specific dates of service restrictions, we could not compare health status or utilization during and outside periods of service restriction. Finally, since data collection was limited to one primary care group and Hamilton-based hospitals, care accessed outside of these locations was not accounted for in our analysis.

## Conclusion

This exploratory research project used parallel convergent mixed methods to describe the healthcare utilization, morbidity, and experiences of shelter users who are service restricted. It describes the relationship between service restriction and health, and underscores potential strategies to improve community health among people deprived of housing, particularly PWUD. We found that people experiencing service restrictions have high rates of primary care, emergency department, and inpatient healthcare use, high rates of patient-initiated discharge in hospital settings, and commonly access care for infections, substance use disorders, wounds, and traumatic injuries. Service restriction and homelessness contributes to the dehumanization of people, leaving them with nowhere to go in the face of opaque and arbitrary service restriction practices. Criminalization is ubiquitous and used to enforce service restrictions, while also deterring people from accessing healthcare. Despite these damaging experiences, people who are service restricted practice care for one another and have several ideas to improve healthcare and social services. In an area with minimal research or evaluation, this study builds the foundation for more robust population-level research on the practice and impacts of emergency shelter service restrictions.

Findings from our study should inform service restriction and shelter policies, including the implementation of safer use spaces in shelters, designated shelters for people seeking sober environments, and access to sterile drug supplies. Service restriction policies should be revised periodically to reflect changes in sociopolitical attitudes, and be transparent to both shelter users and staff. Shelter staff should receive additional training on implementing service restrictions, understanding its impacts, providing written notice of restriction, and assisting those service restricted with finding another shelter. Shelters should also implement a timely appeal process involving an impartial mediator, and systematic data collection on service restrictions to improve transparency and accountability.

Our study has implications for social policy policies at all levels of government. Study participants emphasized that merely accessing shelter is not their goal. Rather, they seek a home and the physical and psychological safety it provides, and housing and homelessness policy should reflect this. Increasing government income supplements (e.g. ODSP payments) is also crucial to affording basic life necessities. From a drug policy perspective, harm reduction and treatment are not opposing forces, but rather complementary approaches that require adequate planning, funding, and resources to ensure PWUD who are deprived of housing can access substance use care and harm reduction options when, where, and how they want to. For health policy, investments are needed in primary prevention along with infectious disease care, preventative care, substance use care, and chronic pain management tailored to people at risk of or experiencing homelessness. From a care delivery perspective, healthcare and shelter staff should receive additional training to provide connected, humanized, and non-judgemental care, supported by proper compensation and accountability systems to sustain this approach. Furthermore, more detailed data tracking in healthcare to identify housing status would enable more robust and generalizable evaluations of these individuals’ healthcare needs.

## Data Availability

All de-identified data produced in the present study are available upon reasonable request to the corresponding authors.

## List of abbreviations

HAMSMaRT: Hamilton Social Medicine Response Team
PWUD: People Who Use Drugs
ERP: Emergency Room Physician
ODSP: Ontario Disability Support Payments
EMR: Electronic Medical Records
EMS: Emergency Medical Services

## Declarations

### Ethics Approval and Consent to Participate

Ethics approval was obtained from the Hamilton Integrated Research Ethics Board [Project #14035]. Our study involves a vulnerable population as per the Tri Council Policy Statement 2, 2018 and we are strongly committed to research practices that respect individual autonomy and protect privacy. Accordingly, we created a private and comfortable environment to conduct interviews, deleted audio files after transcription, only granted EMR access to those conducting chart review, de-identified data, and stored data on an encrypted server in a password protected file. Written consent was obtained from all research participants prior to their participation.

### Availability of Data and Materials

De-identified data used in the current study are available from the corresponding author on reasonable request.

### Author Contributions

SB analyzed data, contributed to knowledge translation, and co-wrote the manuscript. SP contributed to study design, collected and analyzed data, contributed to knowledge translation, and wrote manuscript sections. JP contributed to study design, contributed to participant recruitment, collected and analyzed data, and contributed to knowledge translation. KC contributed to study design and collected and analyzed data. AL contributed to study design and collected and analyzed data. RL collected and analyzed data. OM contributed to study design, collected and analyzed data, and contributed to knowledge translation. AP collected and analyzed data and contributed to knowledge translation. MM contributed to study design, contributed to participant recruitment and analyzed data. FK contributed to study design and analyzed data. LL contributed to study design and analyzed data. RL contributed to study design and analyzed data. TO contributed to study design and analyzed data. CB conceived the project, designed the study, led data collection and analysis, led knowledge translation, and co-wrote the manuscript. All authors edited the manuscript and gave final approval.

## Acknowledgements

The authors express their gratitude to the study participants for sharing their experiences. We’re also grateful for the continued involvement and collaboration of Keeping Six—Hamilton Harm Reduction Action League (https://keepingsix.org/) for aiding us in recruiting study participants. We thank all members of HAMSMaRT (https://www.hamsmart.ca/) for their helpful discussions, and in particular Shabeeh Ahmad for supporting the knowledge translation activities. We also acknowledge previous team members who contributed to earlier stages of our study, including Denene Furman, Gillian Wiwcharuk, Kaley Connelly, and Anjali Sargeant. As a community-based research project, we hold relationships with several participants outside of our study. We were heartbroken and frustrated when one of our research participants died prematurely following their interview for our study. We dedicate this article to their memory and to everyone who died too soon because they were deprived of safe, accessible, and secure housing.

## Authors’ Information

JP, MM, CB, RL, and TO all work together at HAMSMaRT, an organization of health providers and community mobilizers working to integrate clinical practice, critical analysis and socially relevant action. JP and MM have lived experience of being service restricted and housing deprivation, and work in roles where they provide peer support to people who have been service restricted. OM has lived experience of implementing service restrictions as a frontline shelter worker. TO, CB, RL, KC, and AL all provide healthcare to people who have been service restricted.

